# Viral and Host Mediators of Non-Suppressible HIV-1 Viremia

**DOI:** 10.1101/2023.03.30.23287124

**Authors:** Abbas Mohammadi, Behzad Etemad, Xin Zhang, Yijia Li, Gregory J. Bedwell, Radwa Sharaf, Autumn Kittilson, Meghan Melberg, Colline Wong, Jesse Fajnzylber, Daniel P. Worrall, Alex Rosenthal, Hannah Jordan, Nikolaus Jilg, Clarety Kaseke, Francoise Giguel, Xiaodong Lian, Rinki Deo, Elisabeth Gillespie, Rida Chishti, Sara Abrha, Taylor Adams, Abigail Siagian, Peter L. Anderson, Steven G. Deeks, Michael M. Lederman, Sigal Yawetz, Daniel R. Kuritzkes, Mathias D. Lichterfeld, Athe Tsibris, Mary Carrington, Zabrina L. Brumme, Jose R. Castillo-Mancilla, Alan N. Engelman, Gaurav D. Gaiha, Jonathan Z. Li

**Affiliations:** Brigham and Women’s Hospital, Harvard Medical School, Boston, MA, USA; University of Pittsburgh, Pittsburgh, PA, USA; Massachusetts General Hospital, Harvard Medical School, Boston, MA, USA; Dana-Farber Cancer Institute, Harvard Medical School, Boston, MA, USA; Ragon Institute of MGH, MIT, and Harvard, Cambridge, MA 02139, USA; Division of Infectious Diseases, Department of Medicine, University of Colorado Anschutz Medical Campus, Aurora, Colorado, USA; Division of HIV, Infectious Diseases, and Global Medicine, University of California, San Francisco, CA, USA; Center for AIDS Research, Division of Infectious Diseases and HIV Medicine, Department of Medicine, Case Western Reserve University/University Hospitals Cleveland Medical Center, Cleveland, OH, USA; Basic Science Program, Frederick National Laboratory for Cancer Research, National Cancer Institute, Frederick, MD, USA and Laboratory of Integrative Cancer Immunology, Center for Cancer Research, National Cancer Institute, Bethesda, MD, USA; Faculty of Health Sciences, Simon Fraser University, Burnaby, Canada; British Columbia Centre for Excellence in HIV/AIDS, Vancouver, Canada; Division of Gastroenterology, Massachusetts General Hospital, Boston, MA 02114, USA

**Keywords:** HIV, non-suppressible viremia, persistent low-level viremia, producer provirus, integration, CD8 immune response

## Abstract

Non-suppressible HIV-1 viremia (NSV) can occur in persons with HIV despite adherence to combination antiretroviral therapy (ART) and in the absence of significant drug resistance. Here, we show that plasma NSV sequences are comprised primarily of large clones without evidence of viral evolution over time. We defined proviruses that contribute to plasma viremia as “producer”, and those that did not as “non-producer”. Compared to ART-suppressed individuals, NSV participants had a significantly larger producer reservoir. Producer proviruses were enriched in chromosome 19 and in proximity to the activating H3K36me3 epigenetic mark. CD4^+^ cells from NSV participants demonstrated upregulation of anti-apoptotic genes and downregulation of pro-apoptotic and type I/II interferon-related pathways. Furthermore, NSV participants showed no elevation in HIV-specific CD8^+^ cell responses and producer proviruses were enriched for HLA escape mutations. We identified critical host and viral mediators of NSV that represent potential targets to disrupt HIV persistence and promote viral silencing.

## Main

For the majority of persons with HIV (PWH), antiretroviral therapy (ART) suppresses HIV RNA to below the level of commercial assay detection.^1–5^ However, a subset of PWH demonstrate persistent (or non-suppressible) low-level viremia (NSV) while on ART.^6, 7^ NSV has historically been attributed to suboptimal ART adherence and/or accumulating HIV drug resistance.^8, 9^ Previous studies supporting the presence of active viral replication have reported that ART resistance mutations can accumulate when viremia persists in the low but detectable range^10, 11^ and that NSV can increase the risk of virologic failure.^12^ While these factors can cause persistent NSV, other evidence has showed that persistent NSV can be maintained for long periods without leading to high-level virologic failure or the development of new resistance mutations.^13–18^ While suboptimal ART adherence or emerging drug resistance may play a role in a subset of individuals with NSV, alternative mechanisms seem to underlie NSV in other PWH.

Clonal expansion of HIV-infected cells represents a key contributing factor for HIV persistence and recent studies have suggested this plays an important role in NSV as well. Halvas et al. reported that the majority of plasma variants were composed of clusters of identical sequences without signs of active viral replication.^1^ While NSV was fueled by large populations of clonally-expanded HIV-infected cells, the mechanisms that lead to the establishment and maintenance of NSV, and the NSV-generating proviral reservoirs, remain understudied. In this study, we characterized a cohort of eight participants with NSV, and performed in-depth ART drug concentration testing, alongside viral and host cell genetics/genomics and immune profiling. We have identified features of host integration sites that differentiated proviruses fueling NSV from those that were not contributory. Transcriptomic and immunologic phenotyping studies highlighted host cell and cellular immune environments that distinguished PWH with and without NSV.

## Results

### Participant characteristics and assessment of ARV drug levels

We enrolled eight participants, 88% men, with a median age of 60 years and median ART duration of 10 years. The median duration of virologic suppression prior to the NSV and duration of NSV for all participants were 4 and 1.8 years, respectively. During the NSV episodes, the median viral load was 99 copies/ml and the median CD4 count was 798 cells/mm^3^ (Table 1). Individual participant characteristics, ART regimens and genotypic susceptibility scores (GSS)^19^ of plasma viruses sequenced during NSV are shown in Table S1. All participants were receiving at least 2 active antiretroviral drugs during the NSV episodes. Characteristics of the ART-suppressed comparator participants are shown in Table S2 and S3.

We assessed ART adherence by quantifying antiretroviral (and their anabolites) drug concentrations in plasma or through dried blood spot (DBS) testing. LV1 and LV2 had plasma dolutegravir (DTG) and darunavir (DRN) concentrations consistent with ongoing ART use (Table S4). LV3 and LV5-9 had DBS tests for tenofovir (TFV-DP, a measure of cumulative TDF/TAF adherence)^20, 21^ and emtricitabine (FTC-TP, a measure of recent FTC dosing)^22^. The median (range) FTC-TP levels was 5 (4.4-6.7) pmol/punches and TFV-DP levels was 3702 (2771-6684) fmol/punches.^23^ These concentrations are consistent with the highest odds of suppression and lowest odds of future viremia,^24, 25^ suggesting that all study participants should have been virally suppressed on the basis of high adherence. Also, LV3 and LV5-9 had quantifiable FTC-TP, confirming dosing in the preceding 7 days before sampling.^21^ These results demonstrate that our NSV participants had both high levels of short-term and cumulative ART adherence (Table S4).

### Plasma NSV sequences were comprised primarily of large clones without evidence of viral evolution

HIV-1 integration targeting preferences are demarcated by various features of active chromatin, including transcription^26^, histone epigenetic marks^27^, and nuclear speckle proximity.^28^ The provirus landscape morphs over time in response to ART and the host immune response to a quasi-homeostatic state marked by cell loss and clonal expansion.^29–35^ A key goal of this study was to assess aspects of host proviruses that contributed to NSV. Longitudinal single-genome sequencing (SGS) of near-full length proviruses and plasma HIV *pol* and *env* RNA was performed.

A total of 1987 single-genome proviral sequences and 222 single-genome plasma sequences were generated for the 8 NSV participants. Longitudinal plasma HIV sequences were obtained for four participants with available sampling (LV1, LV7, LV8, and LV9), at a median 4.5 time points, an average of 9.7 months apart (Fig. 1a and S1).

**Figure 1.**
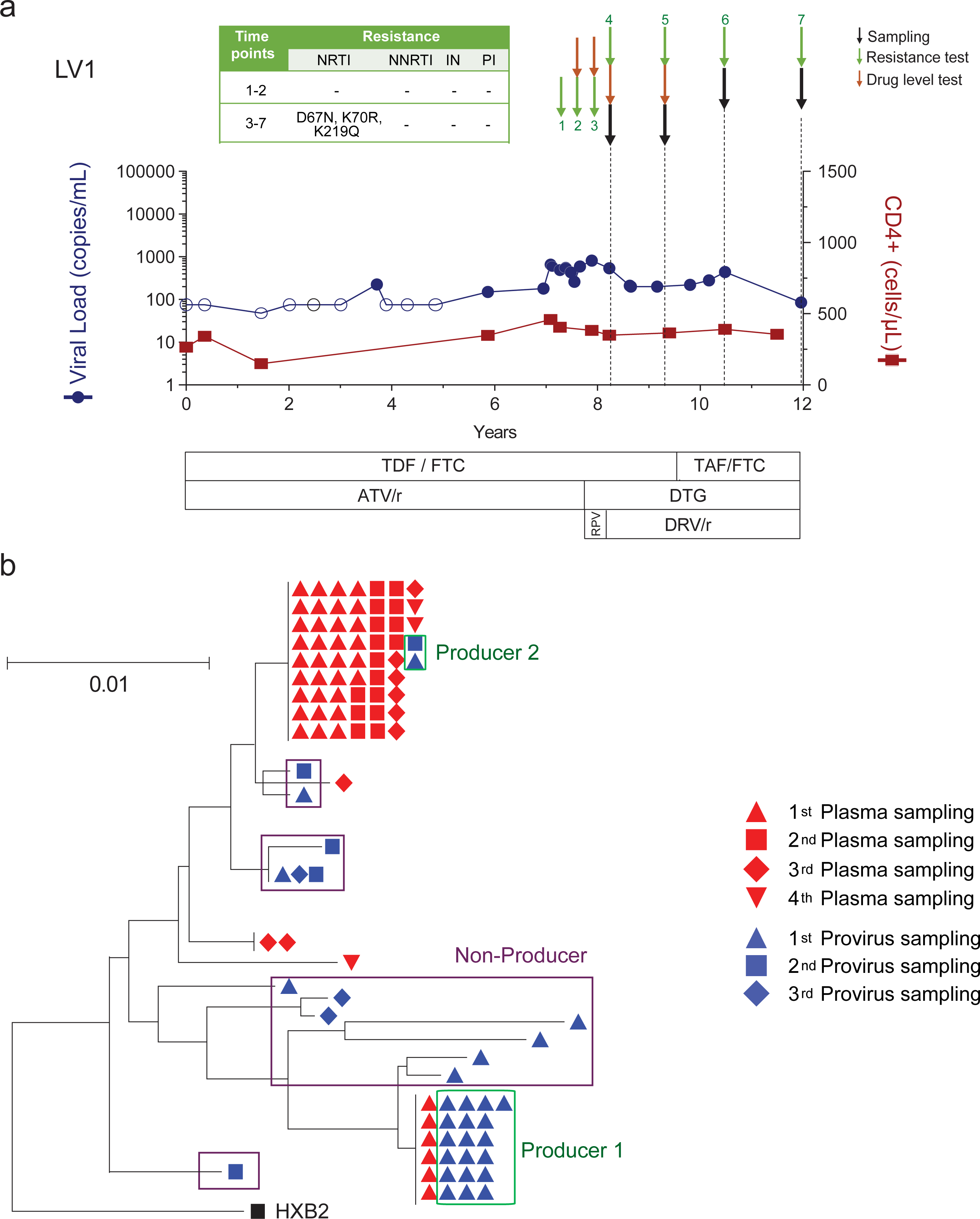
Example participant with non-suppressible viremia (LV1). (a) Viral loads and CD4^+^ T cell count from the time of virologic suppression. Downward green and orange arrows indicate timing of drug resistance and plasma drug level testing, respectively. Sampling times for viral genetic analyses are in black arrows. Antiretroviral resistance mutations are shown in the insert. (b) Neighbor joining trees of proviral and plasma *pol-env* sequences in blue and red, respectively. Producers (green boxes) defined as proviruses with exact matches to plasma RNA sequences. Non-producers (purple boxes) are proviruses that do not match any plasma RNA sequences. Shape indicates sampling time point, corresponding to black arrows in part (a). RPV, rilpivirine; TDF, tenofovir; FTC, emtricitabine; ATV/r, atazanavir/ritonavir; TAF, tenofovir alafenamide; DTG, dolutegravir; DRV/r, darunavir/ritonavir.

Phylogenetic analysis confirmed that sequences from each participant partitioned into separate clusters (Fig. S2). Neighbor joining trees of proviral and plasma sequences for these eight participants showed that the plasma sequences were dominated by one or two clones, with no evidence of viral evolution from longitudinal samplings that would be consistent with active viral replication (Fig. 1b and Fig. 2a). For the initial analysis, proviral sequences were considered intact if they either did not harbor obvious defects or were linked to plasma sequences. At the time of study entry, the 2 largest plasma RNA clones comprised a median 71% (Q1-Q3: 27-83%) of all plasma sequences and were linked to a median 26% (Q1-Q3: 14-61%) of all intact proviral sequences (Fig. S3a). Overall, intact proviruses comprised a median 4.5% (Q1-Q3: 3.8-15%) of the proviral reservoir, with a high degree of variation evident from two participants (LV2 and LV9). LV2 and LV9 proviruses were dominated by several large clones of intact sequences that represented 76% and 34% of their total PBMC proviral reservoirs, respectively (Fig. S3b).

**Figure 2.**
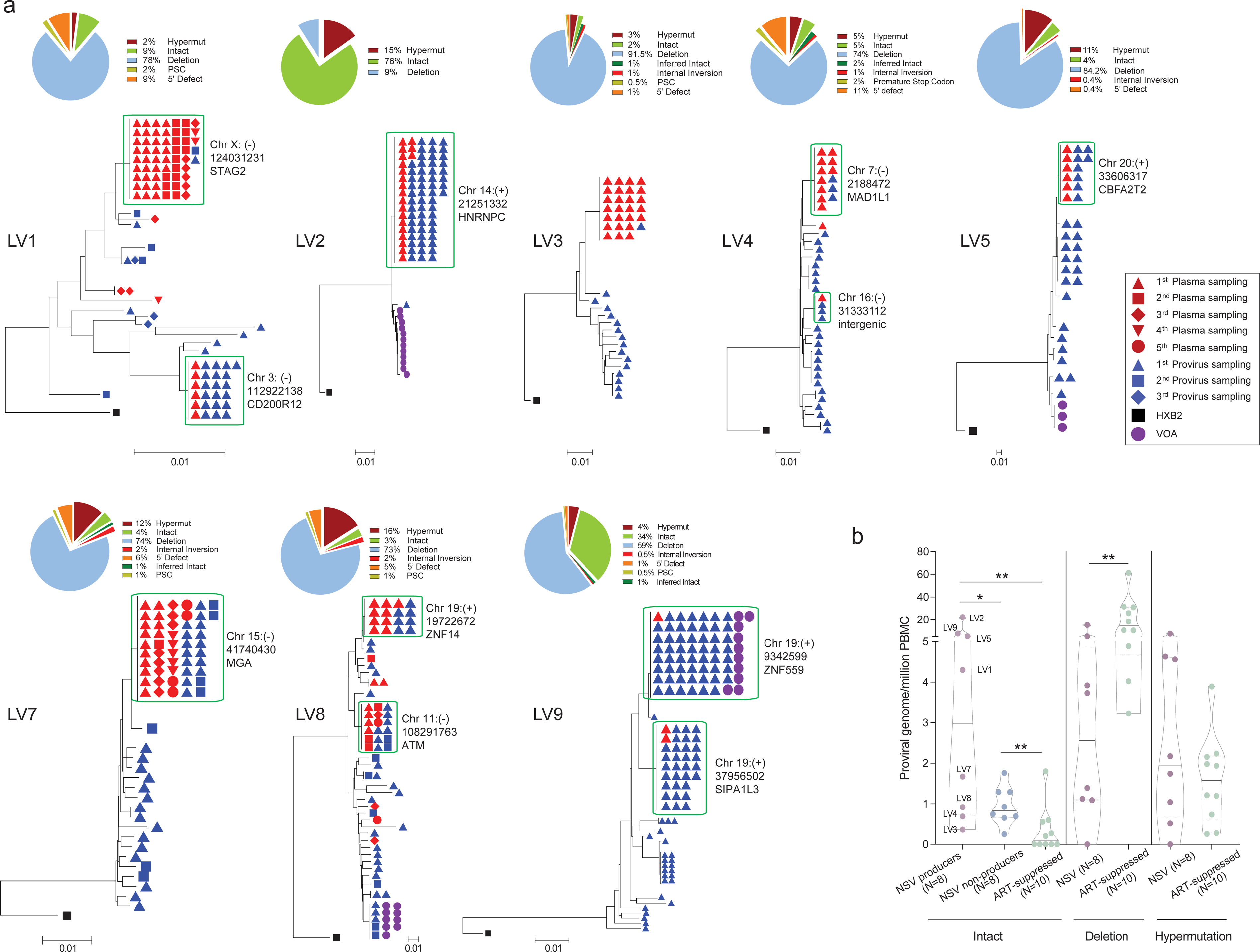
Sequencing overview of the non-suppressible viremia cohort. (a) Pie charts represent percentage of intact and defective proviral sequences for each participant. Neighbor joining trees show intact proviral and plasma sequences from different timepoints. The host integration sites of the producer proviruses are labeled. (b) Comparison of reservoir size (number of proviral sequences per million cells) for intact and defective proviruses between NSV and ART-suppressed individuals. For intact proviruses, a comparison is made of producer proviral versus non-producer proviral reservoir size in NSV participants versus intact proviral reservoir size of ART-suppressed individuals. Wilcoxon matched-pairs signed rank test and Mann-Whitney U tests were used for comparisons. ns, not significant; *P < 0.05, **P < 0.01.

We categorized proviruses as producers if they matched a plasma sequence and as non-producers if they did not. There was a wide range of producer proviruses within the reservoir. For LV2, the PBMC proviral reservoir was largely comprised of one large producer clone representing 98% of intact proviruses, which matched the large plasma NSV clone (Fig. 2a). In contrast, LV3 had the smallest producer reservoir size, representing 3.5% of total intact sequences. These results demonstrate that while these individuals share a common NSV phenotype, their proviral landscape can be highly heterogenous (Fig. S3b).

We next compared the size of the intact and defective reservoir sizes between the NSV participants and a control group of 10 ART-suppressed participants (Table S2). NSV participants had a significantly larger total and intact PBMC proviral reservoir (NSV vs ART-suppressed: median total proviral genomes 34 vs 18 proviruses/million cells, P=0.08 and median intact proviral genomes 4.3 vs 0.1 proviruses/million cells, P=0.001). Specifically, the size of the producer proviral reservoir was significantly larger in the NSV participants than either the non-producer intact proviral reservoir in these participants or the intact proviral reservoir in the ART-suppressed participants (Fig. 2b). In addition, the NSV participants had a smaller number of proviruses with large deletions (median 2.6 vs 10.7 proviruses/million cells, P=0.006). These results suggest that intact reservoir size could be a contributing factor to NSV.

### Integration site and epigenetic signatures of producer proviruses

The location and chromatin landscape of HIV proviral integration sites can modulate the extent of proviral transcriptional activity.^36, 37^ We accordingly evaluated whether certain integration site features differentiated the producer, non-producer and defective proviruses. Using the Matched Integration Site and Proviral Sequencing (MIP-Seq) protocol, we identified host chromosomal integration sites for 11 producer, 21 intact non-producer and 44 defective proviruses across all NSV participants (we were unable to identify an integration site from one LV3 producer clone). Integration sites were identified across all autosomal and sex chromosomes with the exception of chromosome 21 (Fig. 3a). Compared to non-producer and defective proviruses, producer integration sites were enriched in chromosome 19. Twenty-seven percent (3/11) of producer proviruses were located in chromosome 19 compared to none of the 21 non-producer and 44 defective proviruses (producer vs non-producer P=0.03 and producer vs defective P=0.006).

**Figure 3.**
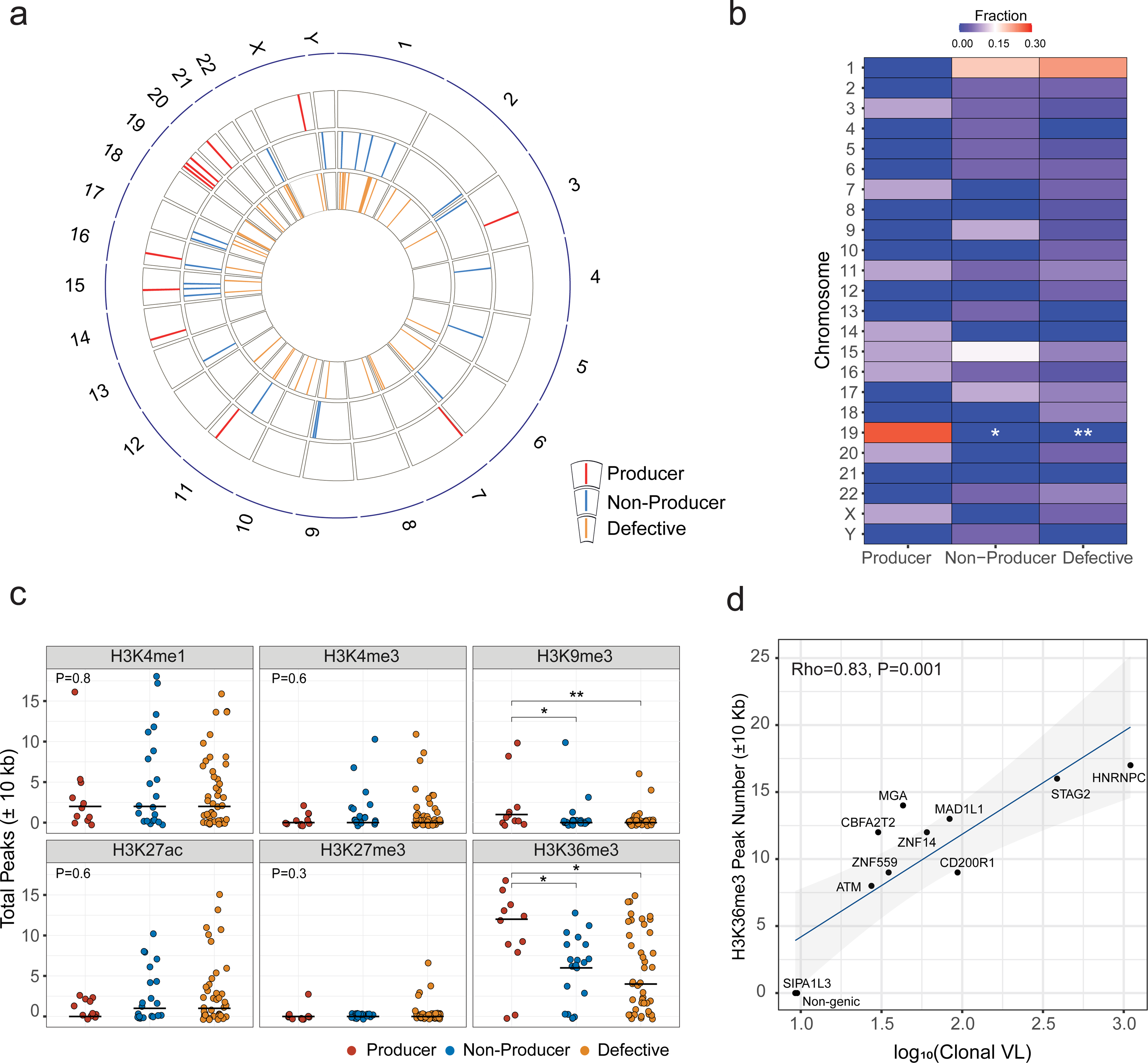
Integration sites and chromatin features of HIV-1 proviruses. (a) Circos plot showing the location of each integration site across human chromosomes. (b) Karyotyping heatmap showing the percentage of integration sites in each human chromosome for different classes of proviruses. Fisher’s exact test was used. (c) Number of peaks for key histone marks in 10 kb regions flanking the proviral integration sites. Mann-Whitney U test was used. (d) Correlation between enrichment of H3K36me3 histone marks near producer proviral integration sites and plasma clone viral loads (viral load multiplied by fraction of plasma sequences matching the producer provirus). Host gene integration sites are labeled. Spearman correlation test was used. *P < 0.05, **P < 0.01.

Significant enrichment of producer proviruses for proximity to two activating epigenetic markers was also observed. Using ChIP-seq data from primary CD4^+^ T cells published on the ROADMAP database,^38^ we detected significantly elevated ChIP-seq reads for the H3K36me3 and H3K9me3 histone marks in proximity to producer integration sites compared to either non-producer or defective integration sites (Fig. 3c). We calculated the level of plasma viral load contributed by the producer provirus, which we call the plasma clone viral load. We observed a significant positive correlation between the number of H3K36me3 ChIP-seq reads in proximity to the producer proviruses integration sites and the plasma clone viral load (Spearman *r*=0.83, p=0.001) (Fig. 3d). Proximity to H3K36me3 has been linked to proviral gene expression^36, 37, 39, 40^, suggesting that producer proviruses are enriched near transcriptionally active regions of the chromosome and that producers could potentially leverage cellular transcriptional machinery for proviral expression and virion production.^36^ In addition, a higher number of proximal ChIP-seq peak numbers for two other activating histone marks (H3K27ac and H3K4me1) were linked to greater expression of host genes containing integrated proviruses (Fig. S4a), although these histone marks were not enriched near producer proviruses.

There were a number of chromosomal features that did not associate with the producer cell proviral phenotype. Distance to transcriptional start sites (TSSs) was statistically indistinguishable between producer, non-producer and defective proviruses, regardless of the orientation of the host gene and provirus (Fig. S4b-d). We also did not detect any significant differences between producer, non-producer and defective proviral classes and their distance to heterochromatic centromeres or the fraction of integration into transcriptionally active speckle-associated domains (Fig. S5a-b).

Finally, using NSV participant CD4 cellular RNA sequencing (RNA-Seq), we found no significant differences in host gene transcript levels between producer, non-producer and defective proviruses regardless of the integration orientation (Fig. S5c-d).

### NSV association with upregulated cell survival signaling and downregulated interferon signaling

Cell survival signaling has been linked to HIV persistence, especially in latently infected CD4^+^ T cells.^41, 42^ To understand the association between cell signaling and NSV, we compared the CD4^+^ T cell transcriptomic features between the NSV group (N=8) and a subgroup of the ART-suppressed individuals (N=5) using RNA-Seq. Compared to the ART-suppressed individuals, NSV participants had 481 upregulated genes and 558 downregulated genes (adjusted P value [Padj]<0.1) (Fig. 4a, red and blue dots). Among these differentially expressed genes (DEG), Gene Set Enrichment Analysis (GSEA) revealed enrichment of pathways related to HIV infection, HIV life cycle, and transcription in the NSV group (Fig. 4b). CD4^+^ T cells from the NSV group exhibited enrichment in oxidative phosphorylation and apoptosis-related signals (Fig. 4b upper panel and Fig. S6a). Specifically, CD4^+^ T cells from NSV participants appeared to be primed for survival via down-regulation of pro-apoptotic genes and upregulation of genes associated with anti-apoptotic pathways, including proteosome-related genes (e.g. PSMB1, PSMB2, PSMD14), ubiquitination-related genes, and oncogenes such as PIK3CA and PIK3R1(Fig. 4c and 4d).^43–46^

**Figure 4.**
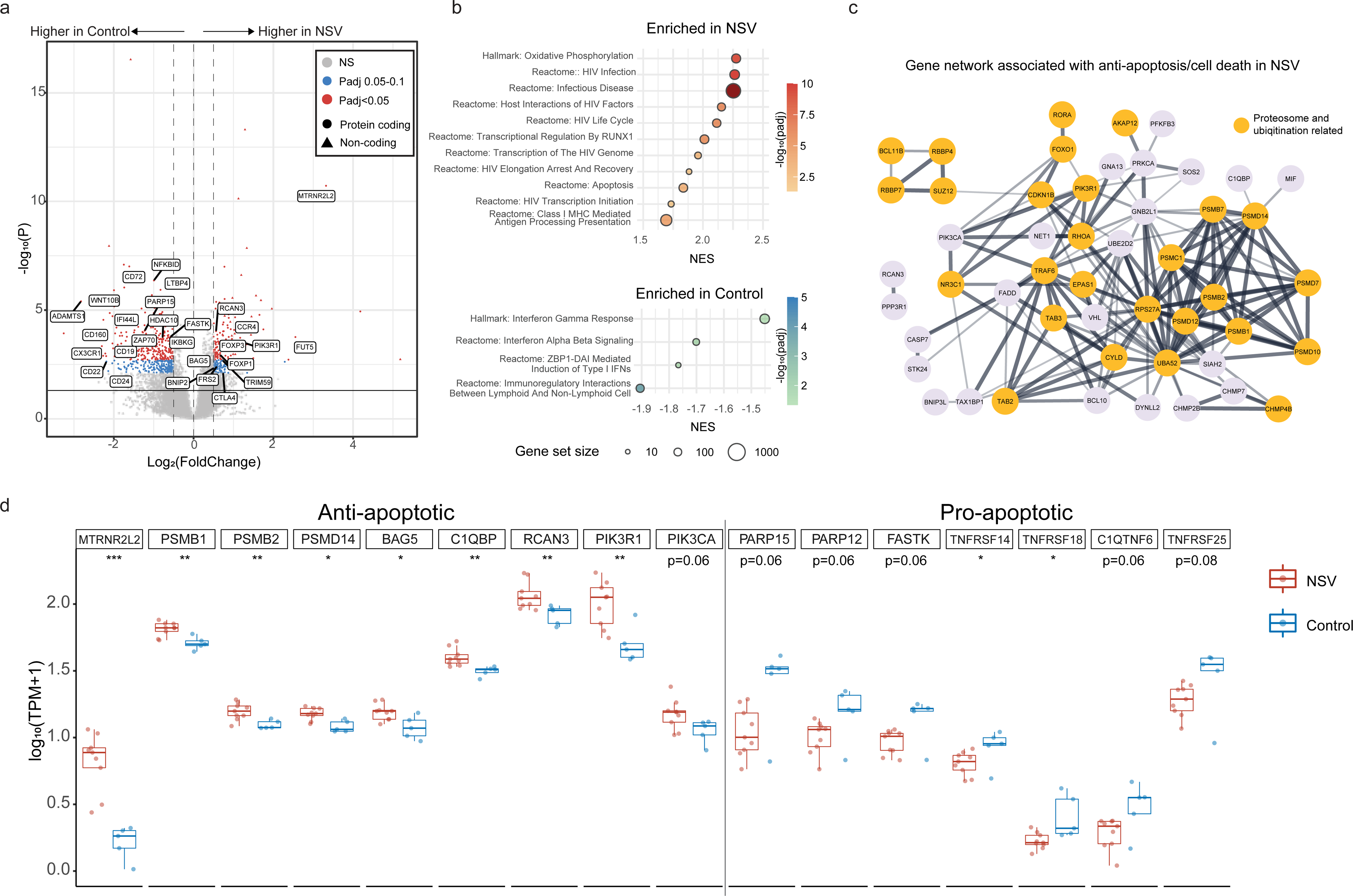
Transcriptomic analysis of CD4^+^ T cells from NSV participants. (a) Volcano plot shows differentially expressed genes in NSV versus ART-suppressed individuals. Red and blue colors highlight different extents of statistical significance. (b) Normalized enrichment score (NES) reflects the degree to which a set of genes is overrepresented among genes that are differentially expressed between NSV and ART-suppressed control participants. Bar-plot represents positively (red) and negatively (blue) correlated pathways. (c) Genes related to proteosome/ubiquitination in NSV participants. (d) Comparing anti-apoptotic and pro-apoptotic gene transcription levels between NSV and ART-suppressed control group. Mann-Whitney U tests were used for comparisons. *P < 0.05, **P < 0.01, ***P < 0.001.

Transcriptomic analysis also highlighted differences in the immune responses between NSV and ART-suppressed individuals. NSV participants demonstrated upregulation of immunosuppression-related genes, including CTLA4 and FOXP3 (Fig. 4a), pointing to an enrichment of the RUNX1-related pathway, which is associated with attenuation in antiviral and interferon (IFN) signaling through FOXP3 binding.^47, 48^ In fact, both IFN-alpha/beta and IFN-gamma signaling were enriched in ART-suppressed individuals (Fig. 4b lower panel, Fig. S6a-c).

IFN signaling plays a pivotal role in HIV pathogenesis by inducing viral restriction factors, causing depletion of CD4^+^ T cells, and regulating systemic immune activation.^49^ These results may point towards potential defects in immune-mediated control of a highly active HIV reservoir as a contributing factor for NSV. Finally, using the random forest algorithm, we identified genes that correlated with the proportion of intact and hypermutated sequences (Fig. S6d). String analysis revealed that the AKT1-centered signaling pathway gene set correlated with the size of the intact proviral reservoir (Fig. S6e).

### Non-suppressible viremia does not increase HIV-specific CD8^+^ T cell responses and is associated with HLA escape mutations

Survival of CD4^+^ T cells harboring producer proviruses not only relies on downregulation of apoptosis and IFN programs, but also resistance to killing.^50^ HIV-specific CD8^+^ T cells, which recognize viral peptides presented in complex with HLA Class-I (HLA-A, -B, and -C), are thought to be one of the most important mediators of viral control.^51^ This appears to be the case even in the setting of ART, as CD8-depletion in the nonhuman primate model leads to loss of viral suppression^52^. Despite the higher antigen-exposure in NSV, we did not detect a more active effector HIV-specific T cell response as determined by IFN-gamma ELISOPT (Fig. 5a). We found no significant differences in HIV-specific CD8^+^ T cell reactivity or proliferation between the NSV and ART-suppressed populations (Fig. 5a and 5b).

**Figure 5.**
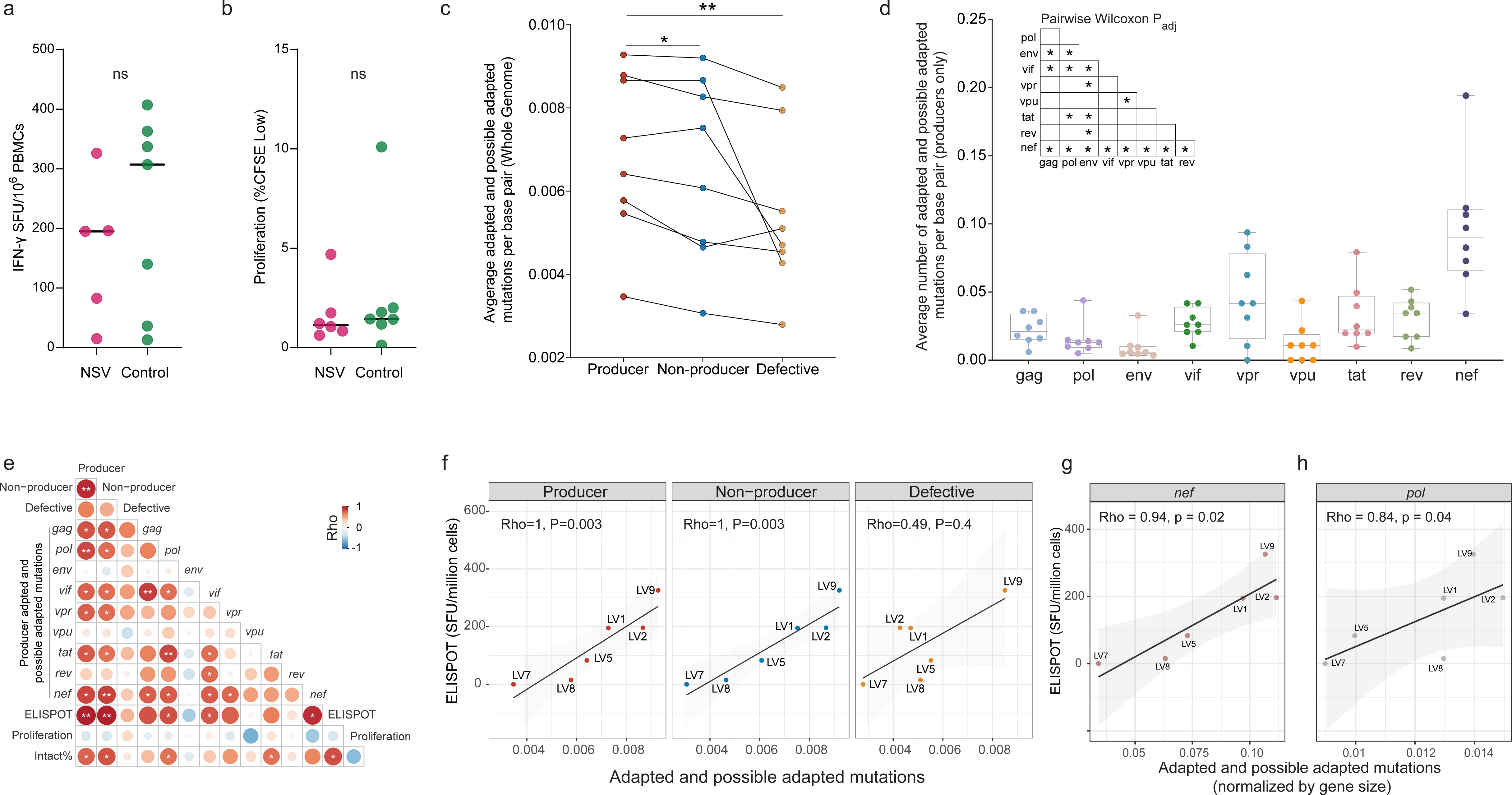
HIV-specific CD8^+^ T cell response and HLA class I escape mutations. (a) HIV-specific CD8^+^ T cell ELISPOT responses in NSV and control (ART-suppressed) cohorts. Mann-Whitney U test was used. (b) HIV-specific CD8^+^ T cell proliferation responses. Mann-Whitney U test was used. (c) Average number of adapted and possible adapted HLA escape mutations across producer, non-producer, and defective proviral sequences. Wilcoxon matched-pairs signed rank testing was used. (d) Average number of mutations per base pair for each HIV gene in intact producer proviruses. Wilcoxon matched-pairs signed rank test was used. (e) Correlation between adapted and possible adapted mutations in different HIV genes in producer proviruses alongside CD8^+^ T cell proliferation activity and percent intact provirus. Spearman correlation test was used. (f) Correlation between adapted and possible adapted mutations in three proviral classes and CD8^+^ T cell activity (ELISPOT). Spearman correlation test was used. (g) and (h) Correlation between CD8^+^ T cell activity (ELISPOT) versus average adapted and possible adapted mutations in *nef* and *pol* in producer proviruses (normalized for gene size). Spearman correlation test was used. ns, not significant; P > 0.05, *P < 0.05, **P < 0.01.

This relatively non-elevated CD8^+^ T cell response in NSV was paired with high-levels of HLA-escape mutations in the producer proviruses. HLA class-I escape has long been recognized as a viral defense mechanism to evade host immune control.^53^ We observed a modestly higher average number of HLA-adapted (i.e., escape) mutations in producer proviruses compared with non-producer (p=0.04) and a dramatically higher escape burden compared to defective proviruses adjusted for proviral length (p=0.001) (Fig. 5c). After normalizing for the size of each HIV gene, *nef* showed significantly higher numbers of adapted and possible adapted mutations compared with other HIV genes (Fig. 5d, S7). Adapted and possible adapted mutations in HIV genomes in both producers and non-producers highly correlated with CD8^+^ T cell IFN-γ release but not with proliferation (Fig. 5e and 5f), suggesting a relationship between effector HIV-specific CD8^+^ T cell responses and subsequent emergence of mutations within proviral clones. Among all genes, the number of *nef* adapted and possible adapted mutations in producer proviruses was strongly correlated with CD8^+^ T cell IFN-γ release in NSV (r=0.94, p=0.02) (Fig. 5e and 5g). Adapted and possible adapted mutations in *pol* in producer proviruses also significantly correlated with total CD8^+^ T cell activity (r=0.84, p=0.04) (Fig. 5h). This seemingly represents immune-driven viral escape mutations that accumulated prior to ART initiation.

### Replication incompetent producers with 5’-deletions in PSI (Ψ) element

Our sequencing revealed that NSV is largely comprised of one or two clonal populations that remain stable over time, which is consistent with high-level viral production from a large, clonally-expanded population of HIV-infected cells as the primary driver of NSV, rather than ongoing viral replication on ART. Thus, producer proviruses driving NSV need not be replication-competent, and we accordingly evaluated producer proviruses for potential replication defects, including deletions in the 5’ PSI packaging element.^54^ In 38% (3/8) of NSV participants, we observed that producer proviruses harbored deletions in the 5’ end of HIV genome (Fig. S8, black boxes). These deletions, which encompassed 22, 15 and 41 nucleotides in participants LV4, LV7 and LV8, respectively, all occurred within SL1 and SL2 elements, ending at the same location within the splice donor site (Fig. S8). Plasma RNA sequencing of the 5’ leader/*gag* region of HIV was performed to confirm the presence of these 5’ defects within the plasma RNA sequences. To evaluate whether these proviruses were infectious, viral outgrowth assays (VOAs) were performed using a transwell system with participant CD4^+^ T cells (LV4, LV7 and LV8; LV2, LV5 and LV9 served as controls) in the bottom chamber and MOLT-4/CCR5 cells in the upper chamber. HIV DNA from the MOLT-4 cells was extracted and subjected to MIP-seq analysis. Producer provirus was isolated from the VOA for LV9 (Fig. 2a and Fig. S9). Non-producer proviruses were isolated from the VOA for LV2, LV5 and LV8 (Fig. 2a).

## Discussion

In this study, we have conducted a comprehensive assessment of NSV and have provided insight into ART-independent factors implicated in HIV suppression and persistence. Our results indicate that suboptimal ART adherence and drug resistance do not appear to be the drivers of non-suppressible viremia. In these participants, NSV is driven instead by the critical intersection of viral and host immune factors. Specifically, the NSV phenotype was highlighted by the presence of large, clonally-expanded reservoirs of proviruses frequently harboring immune escape mutations (and/or defects in the 5’ leader region), integrated in transcriptionally-permissive chromosomal regions, within a CD4^+^ T cell environment primed for survival, and without noticeable HIV-specific T cell responses (Fig. S10).

In one of the first in-depth reservoir studies of NSVs, Halvas et al. reported that NSV is comprised largely of identical populations of plasma viruses that arise from the expansion of HIV-infected CD4^+^ T cell clones, which they termed repliclones.^1^ However, most prior studies sequenced relatively short fragments of viral RNA, which can over-estimate the clonality of plasma sequences. Our plasma RNA sequencing assay combined an ultrasensitive RNA extraction process for 6.7 kb *pol-env* RNA sequencing (Fig. S9a-b). These results confirm that in our cohort, NSV is composed primarily of 1-2 large plasma viral clones that comprised >70% of plasma viruses. The role of these viral clones as the primary driver of NSV was confirmed across multiple longitudinal time points, which failed to reveal evidence of plasma viral sequence changes and evolution. For all of the NSV participants, we were able to identify exact proviral sequence matches for the large plasma clones. We found that the size of the producer proviral reservoir was significantly larger than either the size of the nonproducer proviruses in NSV participants or intact proviruses in ART-suppressed participants, although admittedly with a broad distribution in size of the producer proviral reservoir. The large size of these producer proviral reservoir and their ability to maintain NSV over years highlight the relative stability of this reservoir. These results and the presence of NSV over many years suggests an intrinsic ability of these HIV infected cells to maintain prolonged survival and/or proliferate. Prior studies have reported that CD4^+^ T cells modulating key pro- and anti-apoptotic pathways can maintain survival of HIV-infected cells, drive clonal expansion, and guard against CTLs.^42, 55^ Compared to ART-suppressed participants, CD4^+^ T cells in NSV participants demonstrated transcriptional upregulation of anti-apoptotic pathways and down-regulation of pro-apoptotic pathways. While transcriptional analysis was not isolated to producer cells alone, these results suggest the CD4^+^ T cell environment in NSV participants is primed for survival.

Using the MIP-Seq assay,^39^ we were able to identify the location of HIV integration sites in host chromosomes for 11 producer, 21 intact non-producer and 44 defective proviruses across NSV participants (except producer proviruses from LV3). HIV integration is known to favor active chromatin, and proximity to activating epigenetic marks can modulate proviral gene expression and hence the fate of the resident provirus and infected cell.^56, 57^ We identified certain integration site features that were distinctive in producer proviruses, including enrichment in chromosome 19, which is distinctively enriched for gene density.^58, 59^ We also demonstrated that producer proviruses were located in regions enriched in certain epigenetic characteristics, including a greater number of H3K36me3 histone peaks, which are associated with a transcriptionally-permissive chromosomal regions and elevated proviral expression.^37^ A higher number of H3K36me3 peaks surrounding the producer provirus was also strongly associated with higher plasma viral load comprised of that clone.

Given the transcriptionally-active nature of the producer proviruses, it is unclear why they are not rapidly targeted and cleared by the host immune response. We identified several potential mechanisms that may lead to a more muted immune response to the producer proviruses, including down-regulation of interferon response genes, presence of HLA escape mutations, and lack of increase in activation of HIV-specific CD8^+^ T cell responses despite prolonged elevated levels of HIV antigenemia in NSV. IFN plays a vital role in the innate host antiviral response and contributes to the suppression of HIV viremia.^60^ Compared to ART-suppressed participants, NSV participants significantly down-regulated transcription of IFN genes and multiple genes involved in the IFN-response pathway (Fig. 4), including key regulators IRF3 and IRF7 (Fig. S6b).^61^ Heightened HIV-specific CD8^+^ T cell responses occur with HIV viremia, which is critical to suppress the HIV reservoir.^62^ Thus, we were surprised to find a relatively muted CD8^+^ T cell response, with no significant differences noted in HIV-specific CD8^+^ T cell activity and proliferation between the NSV and ART-suppressed individuals, although there was a clear correlation between the magnitude of CD8^+^ T cell response and mutational burden, suggesting a potential role for viral escape. Interestingly, the loss of SIV-specific CD8^+^ T cell responses can lead to rebound viremia in non-human primates, even in the presence of ART.^52^ Intact proviruses and producers in particular had have high levels of adaptive HLA escape mutations associated with loss of HIV-specific CD8^+^ T cell-mediated clearance of infected cells.^63^ Of note, there was an enrichment of HLA-escape mutations in *nef*, which may be particularly immunogenic as previous studies have reported that the strength of the Nef-specific T cell activity is linked with the size of the HIV reservoir.^64^ Additional studies of Nef function could assess its ability to downregulate HLA-A and B from the infected cell surface, thereby promoting immune escape.^65^

In 38% (3 of 8) of our NSV individuals, we detected deletions in the 5’ leader sequence of the HIV genome. None of these sequences were detected using the VOA, suggesting these proviruses were replication-defective. The 5’-untranslated leader contains several structured motifs that are involved in multiple steps of HIV replication. The deletions are present in the PSI (Ψ) element, which is a highly structured RNA sequence with four hairpin stem loops and a strong affinity for the nucleocapsid (NC) domain of the viral Gag protein. Genome packaging during virus assembly and reverse transcription during the subsequent round of infection are some known functions of the 5’ leader region.^66, 67^ A recent study by White et al. described 4 NSV participants with apparent defects in the 5’ leader sequence.^68^ These defects generally spanned the major splice donor site (MSD) site and resulted in the creation of non-functional virions lacking the envelope glycoprotein. Interestingly, the 5’ leader sequence deletions in our NSV participants spanned the same region and, in fact, LV4 shares the same 22 base deletion that was detected in three participants by White et al. The detection of 5’ leader defects at high frequencies across multiple cohorts suggests a selective advantage of these proviruses in conferring the NSV phenotype, potentially by maintaining a plasma viral load in the absence of HIV replication and/or the ability of Env-deleted virions to escape from host immune surveillance.^69^

In addition to the previously noted limitations, we estimated proviral reservoir size by near-full length proviral sequencing. This could lead to some underestimation of the actual reservoir size, although other methods for reservoir quantification (e.g., IPDA) may over- estimate the size.^70^ Future studies will need to investigate the size of producer proviruses within tissue reservoirs. We found that the peripheral blood reservoir of HIV-infected CD4^+^ T cells contributing to NSV can differ dramatically between NSV participants. It’s possible that NSV- generating CD4^+^ T cells are also distributed within anatomical tissue compartments^71, 72^, especially for those NSV participants with a relatively small producer proviral reservoir size in the peripheral blood. Prior studies have shown that *gag*- or CMV-specific antigens can drive the expansion of certain HIV-infected cellular clones.^31^ Additional studies are needed to delineate which antigens might be playing a role in the expansion of the producer proviruses. Neutralizing antibody responses can also suppress viremia^73^ and evaluation of the humoral immune responses are indicated, although prior studies suggest that some NSV is resistant to autologous neutralizing antibodies.^68^

In this study, we identified critical host and viral mediators of NSV that represent potential targets to disrupt HIV persistence and promote viral silencing. Importantly, ultrasensitive HIV viral load assays can detect residual low levels of HIV viremia in the vast majority of PWH, even on apparently suppressive ART.^74^ Previous studies have reported that such residual viremia is largely comprised of drug-sensitive virus^75^ and relatively homogeneous viral populations.^18, 76^ Thus, we believe it is likely that the mechanisms behind NSV that we describe here are present to some extent in most, if not all, of PWH. Achieving an in-depth understanding of the mechanisms behind NSV may provide insight on strategies for HIV reservoir eradication applicable to all PWH.

## Methods

### Participants

We enrolled 8 ART-treated participants with ≥3 HIV-1 RNA levels between 40-1000 copies/mL over 24 months and compared them to a group of ART-suppressed participants with similar demographic and HIV characteristics. A non-suppressible viremia participant enrolled in the HIV Eradication and Latency (HEAL) cohort, a biorepository of Brigham and Women’s Hospital, was included. The NSV samples were taken from different time points enabling us to study these participants longitudinally. The ART-suppressed comparators included 11 participants from the AIDS Clinical Trials Group (ACTG) and 7 participants from the Ragon Institute of MGH, MIT and Harvard. Written informed consent was obtained from all participants.

### ARV drug level testing

For plasma ARV testing, samples were sent to the infectious disease pharmacokinetics lab at the University of Florida. Testing was performed for darunavir and dolutegravir by liquid chromatography with tandem mass spectrometry. For dried blood spot (DBS) ARV testing, 25 mL of whole blood were spotted five times onto Whatman 903 protein saver cards, as previously described.^21^ After spotting, cards were allowed to dry at room temperature for at least three hours (as long as overnight), after which they were stored at -80°C until analyzed. TFV-DP and FTC-TP were quantified from two 7-mm punches extracted with 2 mL of methanol:water to create a lysed cellular matrix using a previously validated method that was adapted and validated for TAF-containing regimens.^77^ The assay was linear and ranged from 25 to 6,000 fmol/sample for TFV-DP and from 0.1–200 pmol/sample for FTC-TP.^21, 77^

### DNA isolation and HIV reservoir quantification

DNA extractions were carried out from PBMCs using the QIAamp DNA Micro Kit (Catalog #56304), and the quantification of DNA was performed with Nanodrop (Applied Biosystems, ThermoFisher). To estimate the size of the reservoir, we employed NFL-seq, which is described below.

### Near-full length proviral sequencing, sequence alignments, quality control, and Neighbor joining analyses

Extracted DNA was endpoint-diluted and subjected to NFL-seq, as previously described.^78^ We classified our sequences into intact and different classes of defectives (e.g., 5’-defect, deletion, hypermutation, inversion) using a published proviral intactness pipeline.^79^ Proviral sequences were categorized into intact and defective as previously described.^79^ Briefly, after aligning to HXB2, we called our sequences to large deleterious deletions if they have <8000bp of the amplicon size, out-of-frame indels, premature/lethal stop codons, internal inversions, or packaging signal deletions (≥15 bp). If a sequence that was almost full-length exhibited a mapped deletion at the 5′ end, which eliminated the site where the primer binds, but did not display any fatal defects in its sequence, the absent 5′ sequence was deduced to be present, and this sequence was regarded as an "inferred intact" HIV-1 sequence. The Los Alamos National Laboratory (LANL) HIV Sequence Database Hypermut 2. program was used to identify the existence or nonexistence of hypermutations linked to APOBEC-3G/3F. Sequences of the virus that did not have any of the mutations listed earlier were categorized as "genome-intact" sequences. Using MAFFT v7.2.0, we aligned the sequences and utilized MEGA 6 to deduce Neighbor joining trees. We called those intact proviruses with an exact match with plasma sequences as “producers” and other intact proviruses as “non-producers”.

### Plasma RNA sequencing

We sequenced plasma HIV RNA as previously described.^80^ Extracted RNA was diluted to single viral genome levels to meet the criteria of single genome sequencing (SGS) of having no more than one template in each well, theoretically no more than 25% of wells being positive for HIV. Primers were designed to amplify *pol-env*, a 6.7 kb region. The amplification reaction was performed using 0.5 µl primers (10 µmol), 1 µl (10 mmol) MgSO4,1 µl (10 mmol) dNTPs, and 1 U Platinum Taq Polymerase (Invitrogen) in 25 µL total volume. PCR conditions consisted of a denaturation step at 94°C for 2 min, followed by 30 cycles of 30 sec at 94°C, 30 sec at 56°C, 90 sec at 68°C and 10 min at 68°C. Products were underwent Illumina barcoded library construction and MiSeq sequencing. Amplicons were assembled using the UltraCycler v1.0 automated de novo sequence assembly to generate a continuous fragment. Plasma sequences that were within 1-2 nucleotides of the near-full length proviral sequence was considered part of the clonal cluster. We counted the total number of plasma sequences in each clone and divided them by all plasma sequences that we had generated. Then we multiplied the ratio with the plasma viral load determine the contribution of each clone for plasma viral load, which we termed the plasma clone viral load.

For each sequence, the genotypic susceptibility scores (GSS) versus the participants’ ART regimen was calculated using the Stanford HIV database drug resistance scoring system. The Stanford HIV database provides a weighted penalty score for the effect of every resistance mutation and antiretroviral medication with 0 if there is no expected effect to 60 for high-level resistance. For each sequence, the estimated level of resistance for each antiretroviral medication (ARV) was determined by adding all of the penalty scores for each of the drug resistance mutations present. The GSS of each ARV was defined as the following: 1 (Stanford penalty score 0-9), 0.75 (Stanford penalty score 10-14), 0.5 (Stanford penalty score 15-29), 0.25 (Stanford penalty score 30-59), and 0 (Stanford penalty score ≥60). The GSS for the sequence was the sum of the GSS for each ARV as part of the participant’s regimen.

### Total RNA transcripts sequencing (RNA-seq)

CD4^+^ T cells were selected from cryopreserved peripheral blood mononuclear cells (PBMC) using EasySep^TM^ Human CD4^+^ T Cell Enrichment Kit (STEMCELL Technologies Inc.). RNA was extracted from selected CD4^+^ T cells with the AllPrep DNA/RNA kit (Qiagen) with subsequent ribosomal RNA depletion RNA reverse transcribed to cDNA library and sequenced by NovaSeq (Illumina). Sequencing results were processed with the VIPER pipeline for alignment, counting, and quality control.^81^ Differentially expressed gene (DEG) analysis was performed with DESeq2 package^82^ and Gene Set Enrichment Analysis (GSEA) with fgsea package using the adaptive multilevel splitting Monte Carlo approach (n=10,000 for simple fgsea in preliminary estimation of P values).^83^

### Integration site (IS) identification and epigenetics

We characterized single proviral genomes along with their matched genomic integration sites by (MIP-Seq).^39^ Briefly, we initiated whole-genome amplification (WGA) by performing multiple displacement amplification (MDA) with phi29 polymerase using the QIAGEN REPLI-g Single Cell Kit, following the manufacturer’s protocol. Afterward, we divided DNA from each sample and carried out proviral sequencing and integration site analysis. We utilized integration site loop amplification (ISLA), which has been previously described, to obtain the integration sites associated with each viral sequence.^84^ One modification that we made to this assay is targeting both the 5’ and 3’ ends of HIV to assess the integration site on both ends and eliminate any potential bias that may arise from analyzing only one end of HIV. To determine the exact location of HIV in the host gene, we used an online tool for trimming integration sites (https://indra.mullins.microbiol.washington.edu/integrationsites/).85 We analyzed our integration sites for various histone marks by utilizing Chromatin Immunoprecipitation Sequencing (ChIP-Seq) datasets from primary CD4+ T cells that were publicly available on the ROADMAP website.^38^ The NIH Roadmap Epigenomics Mapping Consortium produces a public resource of human epigenomic data to catalyze basic biology and disease-oriented research (http://www.roadmapepigenomics.org/). We calculated the total number of peaks of histone marks in a 10kb window from the flanking sides of the integration site and regarded it as the total peak number. To determine the distance between the IS and the nearest transcriptional start site (TSS), we employed "nearestTSS: Find Nearest Transcriptional Start Site," which is a tool developed in R.^86^

### Assessment of HIV-specific CD8^+^ T cell Reactivity

Peripheral blood mononuclear cells (PBMC) were resuspended at 1x106/mL in RPMI supplemented with 10% FBS (R10) and plated 200 µL per well in Immobilon-P 96-well microtiter plates (Millipore) pre-coated with 2 µg/mL anti-IFN-γ (clone DK1, Mabtech). Individual HLA- optimal HIV peptides matched to each subject’s HLA genotype were added at 1 µM and incubated at 37°C overnight. Negative control wells did not receive peptide and positive control wells were treated with 1 µg/mL anti-CD3 (clone OKT3, Biolegend) and 1 µg/mL anti-CD28 (clone CD28.8, Biolegend) antibodies. ELISOPT assay was performed using manufacturer’s protocol with anti-IFN-γ (clone 1-DK1, Mabtech) capture, biotinylated anti-IFN-γ (clone B6-1, Mabtech) detection, Streptavidin-ALP (Mabtech) and AP Conjugated Substrate (BioRad) followed by disinfection with 0.05% Tween-20 (Thermo Fisher) and analysis using S6 Macro Analyzer (CTL Analyzers). Responses greater than 10 spots per well and 3-fold above negative controls were scored as positive.^87, 88^

### Assessment of HIV-specific CD8^+^ T cell Proliferation

PBMCs were stained at 37°C for 20 minutes with 0.5 µM CellTrace CFSE (Thermo Fisher) as per manufacturer’s protocol at 1x106 cells/mL. Staining was quenched with FBS (Sigma), cells were washed twice with R10, resuspended at 1x106/mL and plated 200 µL per well in 96-well round-bottom polystyrene plates (Corning). Individual HIV peptides corresponding to IFN-γ ELISPOT responses for each patient were added at 1 µM and incubated at 37°C for 6 days before flow cytometric assessment. Negative control wells did not receive peptide and positive control wells received 1 µg/mL anti-CD3 (clone OKT3, Biolegend) and anti-CD28 (clone CD28.8, Biolegend) antibodies. On day 6, cells were stained for viability using Live/Dead Violet (Thermo Fisher), AlexaFluor700-anti-CD3 (clone SK7, Biolegend), BUV395-anti-CD8 (clone RPA-T8, BD Biosciences), and APC-pHLA tetramer matching the peptide used for stimulation, then analyzed by flow cytometry (Fig. S11).

### HLA typing and HIV escape mutation data analysis

HLA-A/B/C typing was performed using sequence-specific oligonucleotide probing (PCR-SSOP) and sequence-based typing as previously described.^89^ We excised individual HIV genes from proviral sequences using Gene Cutter (https://www.hiv.lanl.gov/content/sequence/GENE_CUTTER/cutter.html). We then identified polymorphisms within these genes that are known to be associated with one or more host HLA alleles expressed, as defined using a published list of HLA-associated polymorphisms across the HIV subtype B proteome.^90^ For the escape mutation analysis, each HLA-associated viral site was categorized into one of three groups. 1) “Nonadapted” viral sites showed the specific HIV-1 residue predicted to be susceptible to the restricting HLA, 2) “adapted” sites showed the specific HIV-1 residue predicted to confer escape from the restricting HLA, and 3) “possibly-adapted” sites showed any residue other than the “nonadapted” form, supporting it as a possible escape variant.^91^

### Limiting dilution viral outgrowth assay (VOA)

PBMC from participants and donors without HIV were stimulated with IL-2 (100 U/ml) and PHA (1µg/ml) for 72 hours in R20 culture media. Then, we continued the stimulation with only with IL-2 in R20 culture media. VOAs were performed by using CD4^+^ T cells isolated from cryopreserved peripheral blood mononuclear cells (PBMC) using EasySep^TM^ Human CD4^+^ T Cell Enrichment Kit (STEMCELL Technologies Inc.) from participants and healthy donors. Then we used MOLT-4/CCR5 cell lines and co-cultured those for more than 30 days, as reported previously.^92^ We started with 0.1*10^6^ MOLT-4/CCR5 cells and 0.5*10^6^ CD4^+^ T cells from our participants and healthy donor and cultured them in each well of a 24-Transwell ® plates (STEMCELL Technologies Inc.). We collected samples from supernatant and MOLT-4/CCR5 every 3 days and refreshed the media with IL-2 (100 U/ml) in R20.

## Data analysis

We analyzed our results by using Mann-Whitney U tests (2-tailed), Fisher’s exact tests, Wilcoxon’s tests as appropriate. Correlations were tested by the Spearman’s rank test. Adjustment for multiple comparisons was made in the analysis of ChIP-seq histone marks and host gene transcription, RNA-seq, and numbers of HLA escape mutations per HIV gene. A P-value of less than 0.05 was deemed significant. We adjusted for multiple comparisons in the analysis of ChIP-seq histone marks and host gene transcription, RNA-seq, and the number of HLA escape mutations per HIV gene. However, we did not make any corrections for multiple comparisons in the other analyses, as it was an exploratory analysis. We performed the statistical analysis using Prism (GraphPad v.7) and the statistical packages in R (R Project for Statistical Computing, version 4.1.0).

## Study approval

All study participants provided written informed consent. The study was approved by the Mass General Brigham Institutional Review Board.

## Data Availability

All data produced in the present study are available upon reasonable request to the authors

## Acknowledgement

This work was supported in part by the National Institutes of Health (NIH/NIAID) grants AI125109 (to JZL), R37AI039394 (to ANE), U54AI170791 (to ANE), Harvard University Center for AIDS Research (5P30AI060354-08 to JZL, 5P30AI060354-14 to GQL) and a subcontract from UM1AI106701 to the Harvard Virology Support Laboratory (to JZL). The content of this publication does not necessarily reflect the views or policies of the Department of Health and Human Services, nor does mention of trade names, commercial products, or organizations imply endorsement by the U.S. Government. This Research was supported in part by the Intramural Research Program of the NIH, Frederick National Lab, Center for Cancer Research. ZLB is supported by a Scholar Award from Michael Smith Health Research BC. We are grateful for the contributions of the participants who made this study possible. We appreciate the support of the staff at the MGH sequencing core facility. We thank Dr. John J. Szela for help with enrollment, Trevor James Mitsutoshi Tamura for his valuable feedback, Zach Herbert and the Dana Farber Cancer Institute Genomics Core Facility.

## Data availability

All data and code are available by request. Sequence data were submitted to Genbank (Accession numbers *Pending*). ROADMAP epigenomic data are available at http://www.roadmapepigenomics.org.

